# SARS-CoV-2 Pandemic Preventive Methods Efficacy - A Simulation Case Study

**DOI:** 10.1101/2021.10.17.21265111

**Authors:** Malak Saad, Emad M. Boctor

**Affiliations:** Del Norte High School, San Diego, CA; Johns Hopkins University, Departments of Radiology and Computer Science, Baltimore, MD

## Abstract

The world has been facing the SARS-CoV-2, a.k.a. COVID-19, pandemic with different preventive methods including social distancing, face masking, screening tests (a.k.a. active surveillance), and vaccination. There are many publications and studies on the efficacy of each of these preventive methods for the last couple of years. Not all methods are readily available in each country and not all methods are accepted by all people in each society.

In this study, we explore the interaction of the three preventive methods: face masking, vaccinations, and screening tests. We study a confined space to represent schools, businesses, or healthcare facilities and we model the spread of the COVID-19 virus for a 60-day period among a sample population while varying the percentage of people adopting one or more of the three preventive methods.

To interpret the simulation results, we define a (Health Goal) target, for example achieving <5% infection rate, i.e., protecting 95% or more of the sample population. We then construct a (Decision Tree) that depicts all valid combinations that achieve this goal. Multiple scenarios are derived from the decision tree to guide decision makers in drawing effective policies to contain the virus spread. We demonstrate a ramping vaccination rate scenario, a removal of the face-masking mandate scenario, and a cost-minimizing goal scenario.

The study highlights the efficacy of combining the three prevention methods to constrain the virus spread among the sample population. For example, results show that a combination of 0% vaccination rate, 6% daily screening test rate, and 80% face masking rate will achieve the target ≥95 protection rate, which can represent a scenario in which vaccination is not yet readily available. As the vaccination rate ramps up to 80% among the sample population, the screening test rate can be 0%, while the face masking rate can be as low as 5% to still achieve the health target. Many other scenarios are derived from this study to meet the defined health target, which represents the flexibility afforded to policy and decision makers when trying to adopt a combination of these preventive methods to contain virus spread.

The study also reveals the higher efficiency of either the vaccination or screening test over face masking under the assumed virus transmissibility rates in the study.

## Introduction

According to the Johns Hopkins Coronavirus Resource Center^**1**^, as of October 2021, global cases of the COVID-19 pandemic exceeded 240 million people while the death toll exceeded 4.8 million. Since it was first announced in December 2019, the world has been struggling to contain the spread of the virus. Many early actions have been taken such as lock downs of whole countries or cities, stay-at-home mandates, and travel restrictions.

SARS-CoV-2 spreads mainly via respiratory droplets. Respiratory droplets travel into the air with coughing, sneezing, or talking, for example. Social distancing is an effective method for protecting the population from the spread of the virus according to the Centers for Disease Control and Prevention (CDC), who recommend 6-feet distancing^**2**^, and the World Health Organization (WHO), who recommend 1-meter distancing^**3**^. Wearing face masks that cover the mouth and nose is another effective mitigation method recommended by the CDC^**4**^ and WHO^**5**^. Many publications have demonstrated the benefits of masking in reducing the spread of the virus^**6, 7, 8, 9**^. Some researchers have relied on mathematical modeling tools to investigate the efficacy of face masks to prevent the spread of the virus^**10, 11**^.

The SARS-CoV-2 virus testing has now become reliable and available^**12**^, however the testing is not yet completely affordable and available to many countries in the world or many of the low-income populations^**13**^. COVID-19 vaccinations have also become readily available, and some are fully approved by the FDA. More than 188M Americans have been vaccinated as of October 2021^**1**^. Similarly, vaccine availability and affordability in many developing countries around the world is still challenging^**14, 15**^. Many clinical trial publications have proven the efficacy^**16, 17, 18, 19**^ and efficiency^**21, 22, 23**^ of the different vaccines. Many variants of the ancestral viral strain have developed, including Alpha, Beta, Gamma, and Delta. According to the CDC, vaccines currently approved in the USA are highly effective against hospitalization and death for these strains, and early data show fully vaccinated people are less likely than unvaccinated ones to re-acquire or transmit SARS-CoV-2^**24**^.

In this paper we conduct a simulation study for the interplay of the three virus prevention methods: face masks, screening tests, and vaccinations. Since these mitigation methods coexist in each country and community with varying degrees of compliance and availability, it is critical to understand the efficacy of combining them on the health of the population.

## The Simulation Study Design

From the early days of discovering the SARS-CoV-2 virus, there have been many publications on simulating the spread of the virus among populations, one of the earliest of which was the Washington Post article^**25**^ that involved the simulation of virus exponential spread and several suggested ways to flatten the curve using social distancing and mask wearing practices. Last year, our team published a simulation study regarding the health and economic impacts of Active Surveillance (screening tests) in a school environment^**26**^. The simulation study is based on the Coronavirus Simulation Matlab program written by Joshua Gafford^**27**^, which is a recreation of the Washington Post COVID-19 simulation study. The Matlab model simulates SARS-CoV-2 transmission among a sample human population in a confined space. A simple multibody physics model, based on the elastic collision between two equal-mass particles, was implemented to simulate people’s trajectory within the confined space. In the current study we investigate the efficacy of the three prevention methods (face masking, screening tests, and vaccination) on controlling the spread of the virus within that space.

We simulate a population of 500 individuals interacting among themselves daily in a confined space for the duration of 60 days. This simulation can represent a school environment, a healthcare facility, or a business environment, for example. Nowadays, we are witnessing different attempted strategies of government officials, school district officials, and business leaders to transition their employees or students safely to their in-person environment. The different strategies include hybrid models of returning and working from home, masking and social distancing, weekly screening tests, or reporting vaccination status. According to our knowledge, there is no sufficient evidence of a clear winning formula for combining these strategies to achieve the highest rates of protection for the population. Our main goal of the study is to define patterns of combining these prevention methods to achieve a health target, which can provide helpful guidance to decision makers in deciding the optimal ways to control the spread of the virus.

The simulation assumes only 1% of the 500-person population is infected initially with the SARS-CoV-2 virus. The population is spread randomly within the confined space and we simulate their random movements for 60 days with several daily movements. When two individuals come in contact within a certain proximity circle, they interact and possibly transmit the virus from one sick person to another healthy one. We simulate an average recovery time of 14 days for a sick person, and a recovered person will not catch the virus again for the duration of the simulation.

To simulate the interaction of the three prevention methods, we assume a percentage of the population abides by the face mask mandates (the mitigated population), and we assume another percentage of the population is vaccinated (the vaccinated population). The mitigated and vaccinated populations are assumed to be mutually exclusive in this study, so no individual can be both masked and vaccinated. To simulate the screening test method, we simulate a percentage of the population to be randomly selected daily for a screening test that gives immediate results. If any infected person is detected, they are put in quarantine for 14 days. Table1 summarizes the percentage rates used for each prevention method in the simulation.

**Table1.** Virus spread prevention methods percentages used in the simulation

| Prevention Method | Percentages |
| --- | --- |
| **Screening Test percentages (SP)** | 0%, 2%, 4%, 6%, 8%, 10%, 20% |
| **Face Masking percentages (FP)** | 0%, 5%, 10%, 20%, 30%, 40%, 50%, 60%, 70%, 80% |
| **Vaccination percentages (VP)** | 0%, 5%, 10%, 20%, 30%, 40%, 50%, 60%, 70%, 80% |

All the valid combinations of the above percentages are simulated. Some combinations are invalid when the total percentage of the vaccinated population, face masking population, and the 1% infected population exceed 100%. We average each combination ten times and calculate the mean value and normalized standard deviation. The simulation combinations totaled to 504, which results in 5040 simulation runs when each combination is averaged 10 times.

Table2 summarizes the assumed infection transmissibility rates under different conditions of two individuals who come in close contact. The SARS-CoV-2 virus is highly contagious and thus we assumed a baseline infection transmissibility rate of 99% between two individuals who are not taking any preventive action while one of them is infected. The efficacy of the face masks and vaccinations are reported in the literature but with varying degrees. Stutt et. al^**10**^ assumed 50% and 75% mask effectiveness in their simulations. M. van der Sande et al.^**28**^ found cloth mask filtration rates of 60% of particles between 0.02 μm and 1 μm. We assumed 50% transmissibility rate when one individual is wearing a mask and 25% when the two individuals in contact are both wearing masks. The CDC^**24**^ referenced many clinical trials for vaccine effectiveness and many trials proved 90% or more efficiency. We assumed 10% transmissibility between an infected person and a vaccinated one. If two individuals are vaccinated, then we assumed 1% transmissibility. In case of one face masked and one vaccinated person, the transmissibility is assumed to be 5% (50% x 10%.) It is worth noting here that the assumed infection rates in our study are not absolute but rather represent a valid study case to derive health protection patterns and understand the interplay of these three prevention methods. The individuals who are randomly picked daily for screening test are blindly selected among all individuals in the study, either they are vaccinated, face masking, or neither.

**Table2.** Assumed SARS-CoV-2 transmissibility rates

| Simulation Case | Assumed Transmissibility Rate |
| --- | --- |
| Baseline transmissibility rate between an infected person and uninfected person who are neither wearing face masks nor vaccinated | 99% |
| Transmissibility rate between an infected person and uninfected person while only one of them is wearing a face mask | 50%<sup>10</sup> |
| Transmissibility rate between an infected person and uninfected person who is vaccinated | 10%<sup>24</sup> |
| Transmissibility rate between an infected person and uninfected person who are both wearing face masks | 25% |
| Transmissibility rate between an infected person and uninfected person while one of them is wearing a mask while the other is vaccinated | 5% |
| Transmissibility rate between an infected person and uninfected person who are both vaccinated | 1% |

The main outcome of each simulation run is the percentage of people who are “unaffected” by the virus, meaning they avoided getting infected at all during the 60-day simulation period. We also calculated the percentages of infected population and recovered population. We assumed the worst case of asymptomatic or pre-symptomatic disease pattern where sick individuals would still interact with peers and possibly infect others.

### The Simulation Environment

To demonstrate the simulation environment, we show an example of one of the 5040 simulation runs in Figure1. In this run 2% of the population are randomly selected and screened daily for SARS-CoV-2 virus. All individuals who test positive are quarantined for 14 days and thus cannot infect anyone else while recovering. 20% of the population in this case are vaccinated and 30% of the population are wearing masks. The graphical representation on the left of the figure shows dots to represent individuals. The dots are color-coded as follows: green dots are the healthy, unaffected, individuals who did not catch the virus during the 60-day trial, black dots represent vaccinated individuals, red dots represent infected individuals, and blue dots represent individuals who got infected during the trial but then recovered within 14 days. The graphical curve representation on the right side shows the daily update of each population category (unaffected, infected, vaccinated, and recovered.) In this simulation run, the unaffected percentage is about 43% at the end of the trial period, which indicates that 57% of the population got infected during the trial. We conclude from this example that the mixture of 2% screening test, 20% vaccination, and 30% face masking results in protecting 43% of the population from catching the virus.

**Figure1.**
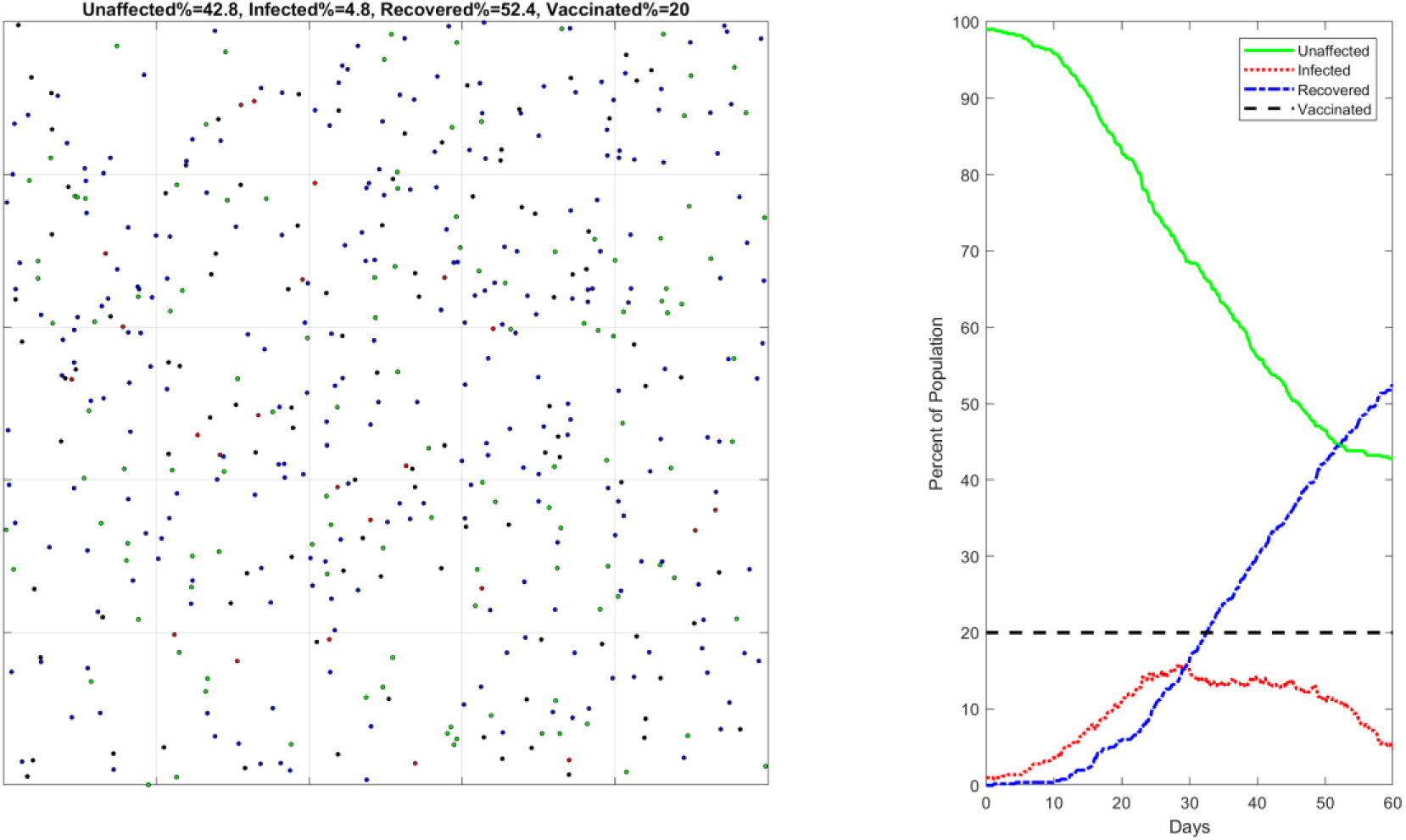
An example simulation run with 20% vaccination percentage, 2% screening test percentage, and 30% face masking. Left: simulation of the population distribution within a confined space. Right: simulation metrics update curves: Unaffected%, Vaccinated%, Infected%, and Recovered%. At the end of the 60-day trial, the unaffected population rate is about 43%, meaning 57% of the population got infected at one point during this trial.

It is worth showing the baseline simulation run in Figure2 where it is assumed that 0% of the population gets tested, vaccinated, or wears face masks, resulting in no prevention methods applied. In this case only 1.6% of the population did not catch the virus by the end of the 60 days while 98.4% got infected.

**Figure2.**
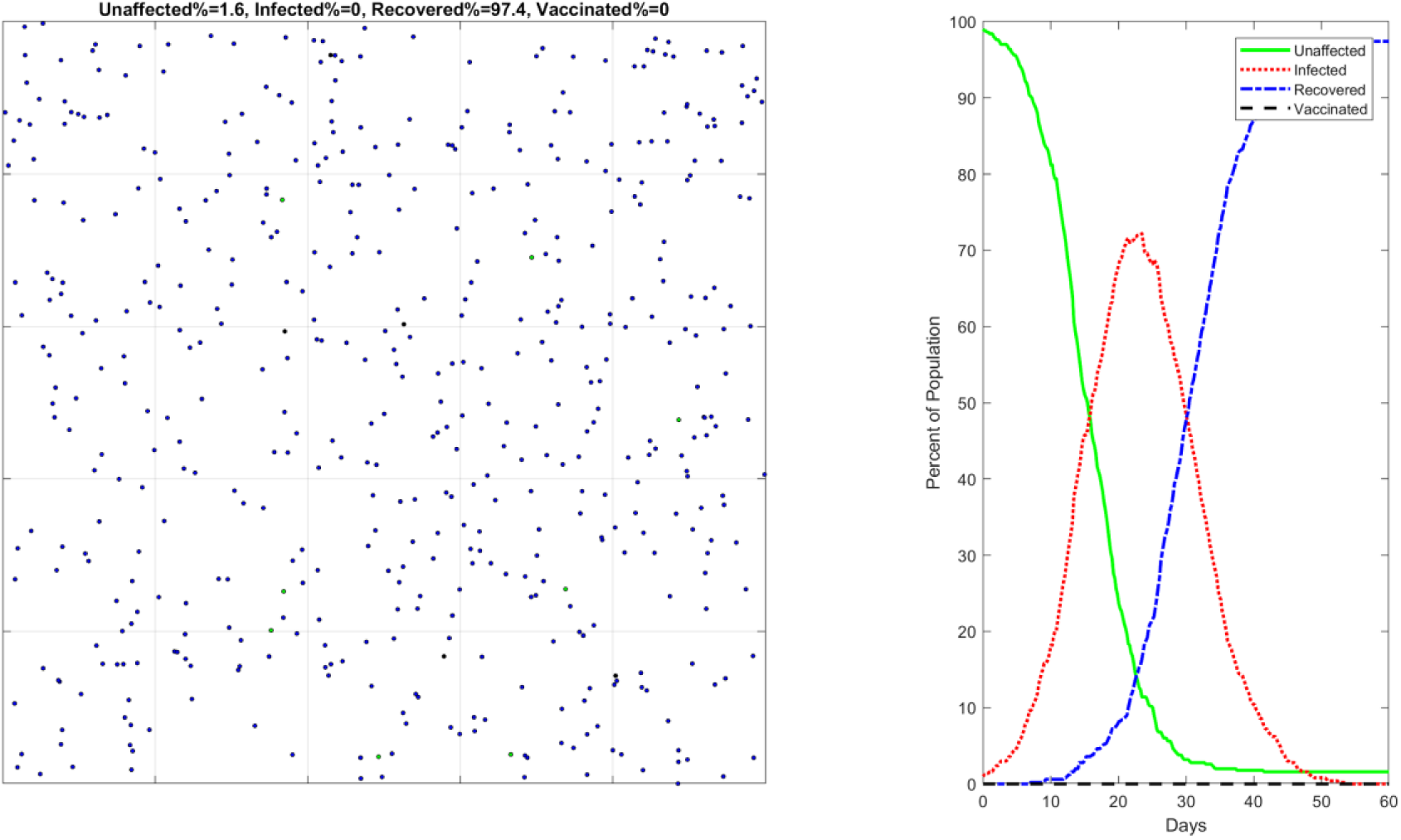
The baseline simulation run with 0% screening test, 0% vaccination, and 0% face masking. Left: simulation of the population distribution within a confined space. Right: simulation metrics update curves: Unaffected%, Vaccinated%, Infected%, and Recovered%. At the end of the 60-day trial, only 1.6% of the population is unaffected by the virus, which concludes that 98.4% of the population got infected.

Figure3 demonstrates another example where all the three prevention methods are improved. 4% of the population is randomly screen tested daily, 40% is vaccinated, and 50% wears masks. It is evident in this case that 95% of the population remains unaffected by the virus (only 5% are infected), which indicates a major improvement in protecting the sample population.

**Figure3.**
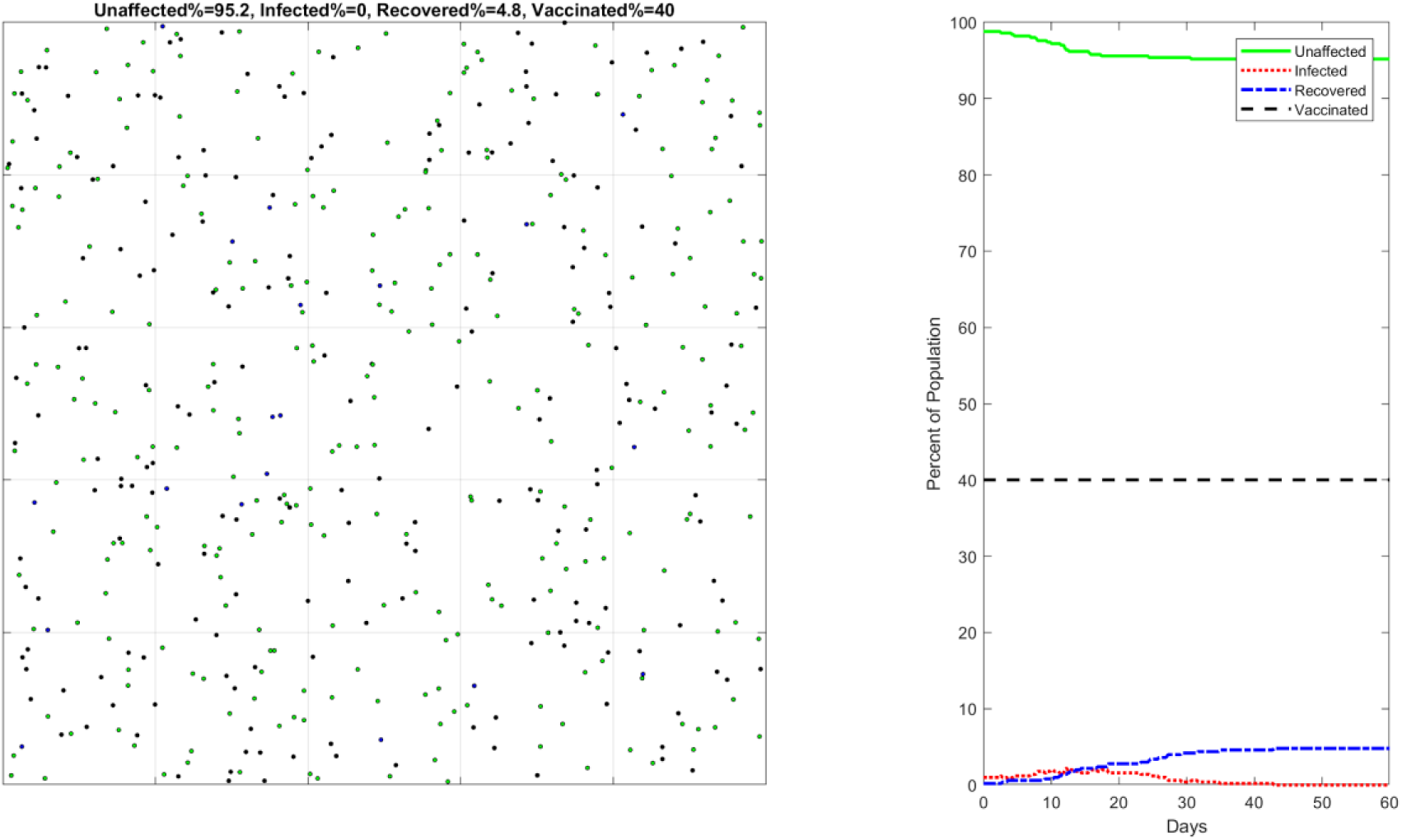
A second example simulation run with 40% vaccination, 4% screening test, and 50% face masking. Left: simulation of the population distribution within a confined space. Right: simulation metrics update curves: Unaffected%, Vaccinated%, Infected%, and Recovered%. At the end of the 60-day trial, the unaffected population rate is about 95%, i.e., the infected population rate is about only 5%.

### Simulation Results and Analysis

The interplay of the three prevention methods listed in Table1 resulted in 504 unique valid combinations, each of which is averaged 10 times to result in a total of 5040 simulation runs. The input to each simulation run is a combination of percentages of the screening test, vaccination, and face masking; the output is the percentage of the unaffected or healthy individuals by the end of the 60-day trial run. The unaffected % is an indication of the infection rate among the population. Figure4 is a graphical representation of the averaged simulation results of each of the 504 prevention methods combinations. The three axes in the graph represent the three prevention method percentages of each run (vaccination %, face masking %, screen test %). Each dot in the 3D scatter graph represents the unaffected population percentage (averaged from 10 runs) and is color coded by its value.

**Figure4.**
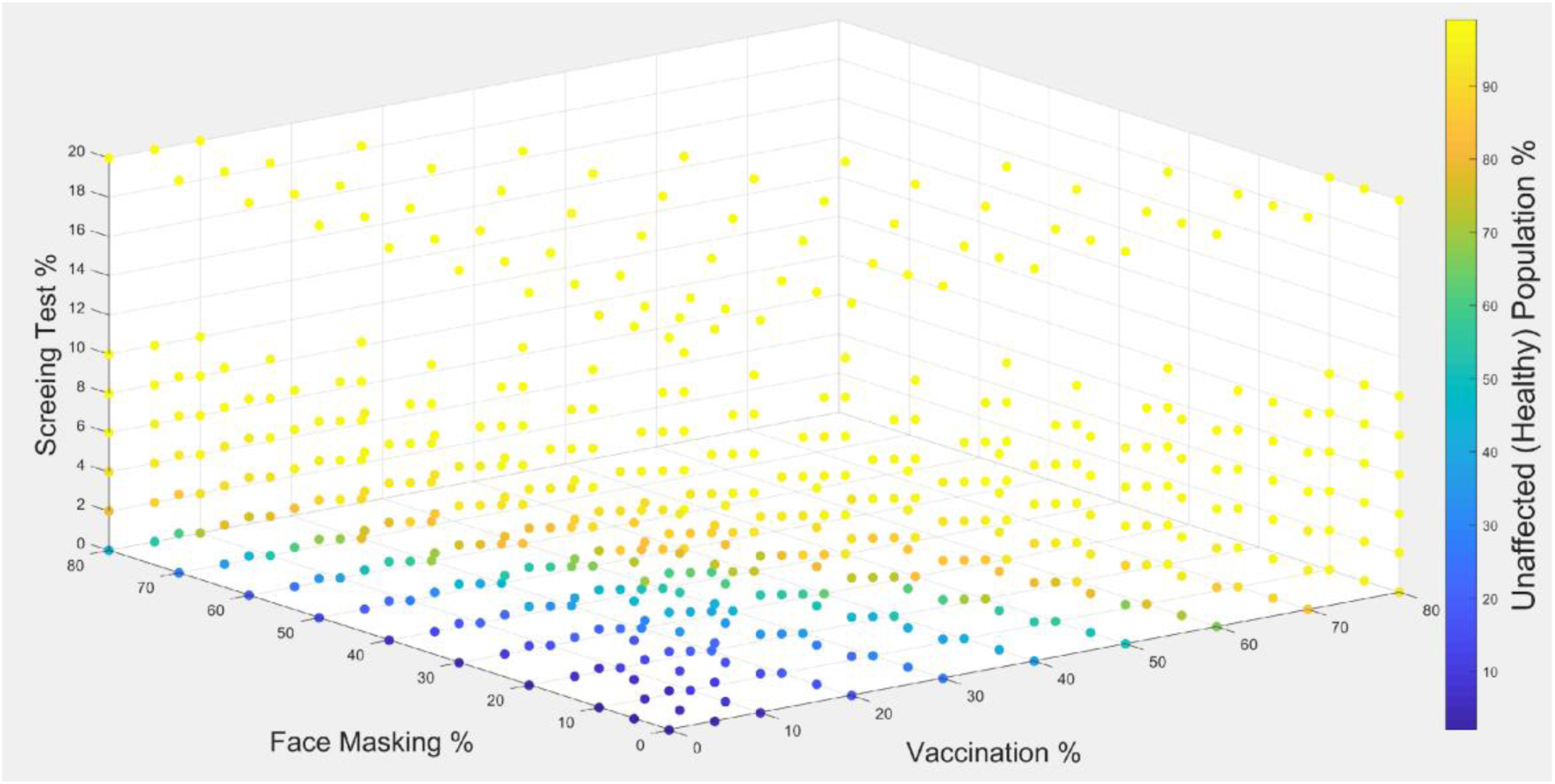
A 4D scatter plot graphical representation for the simulation results. Each dot represents an averaged simulation run with a specific combination of screening test %, vaccination %, and face masking %. The dots are color-coded with the percentage of unaffected (healthy) individuals for each simulation run.

It is evident from the graph in Figure4 that all three prevention methods are effective in reducing the infection rates, and their effectiveness increases as they are combined. It is also clear that smaller percentages are not effective in constraining the infection. Figure5 demonstrates the histogram of the normalized standard deviations (Normalized STDEV = STDEV / Mean) of all 504 averaged simulation runs. The histogram shows that 69% of the simulation runs have normalized STDEV less than 0.1 or 10%, and 98% of all runs have STDEV less than 0.3 or 30%.

**Figure5.**
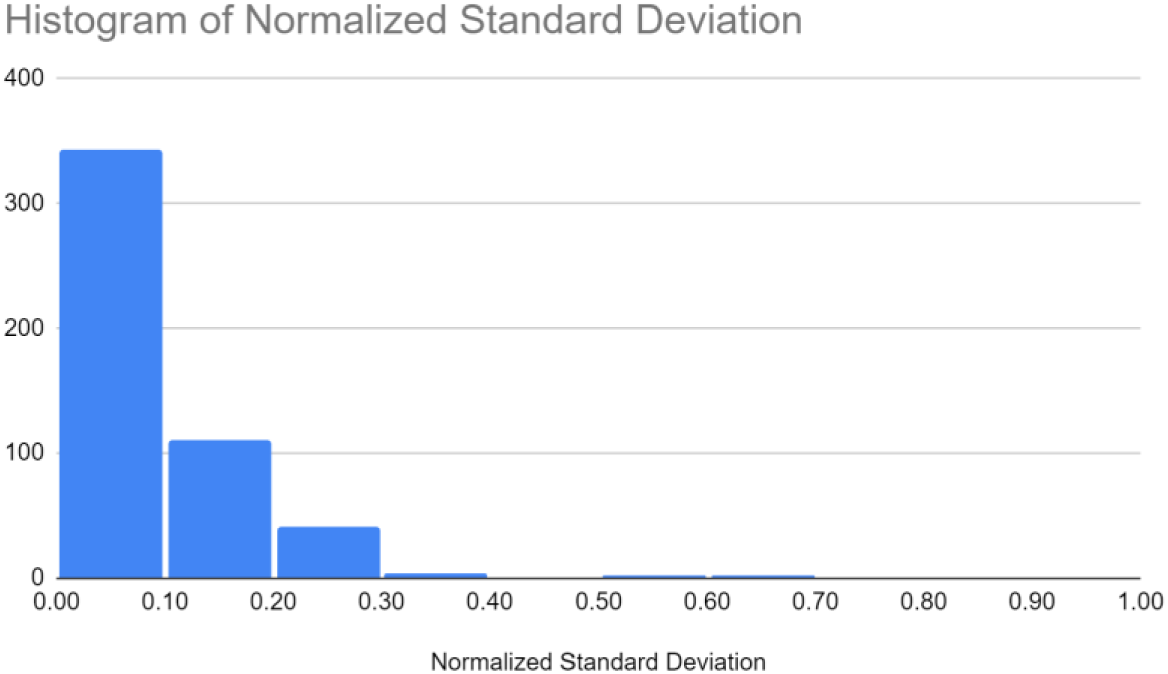
The Normalized Standard Deviation histogram of all simulation runs

The main goal of our simulation study is to understand the interplay between all three virus prevention methods. As an example of how to interpret the simulation results, we defined a health target of achieving less than 5% infection rate among the sample population. We identified all combinations of the three prevention methods that can achieve this target. We used a “Decision Tree” representation to visually present these entries as shown in Figure6 below. The decision tree lists the ramping vaccination percentages from 0% to 80% while listing all associated combinations of screening test and face masking percentages. As seen in the figure, there are many combinations that achieve the <5% infection rate health target.

**Figure6.**
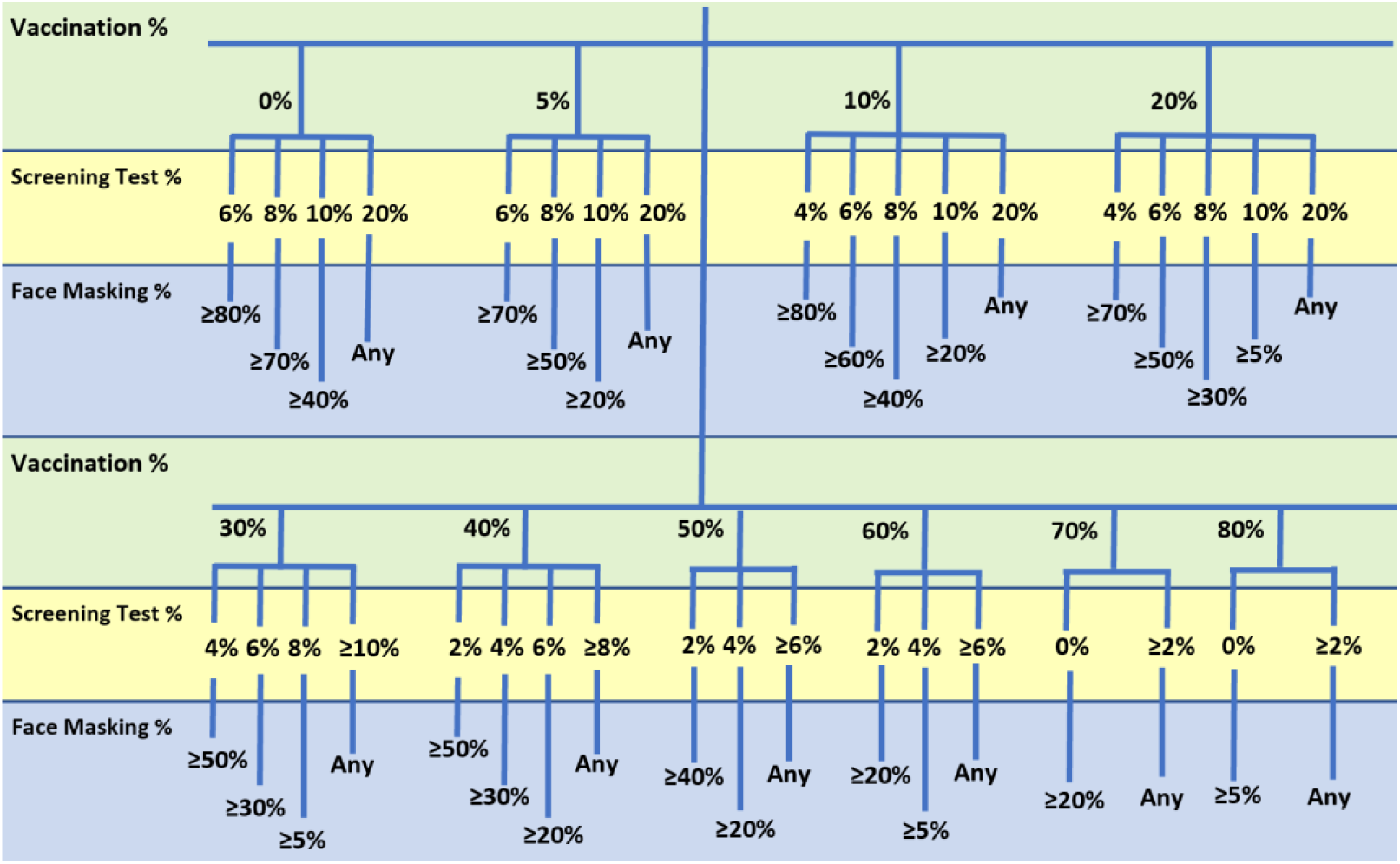
Decision Tree representation for the combinations of all three prevention methods percentages that achieve <5% infection rate (≥95% healthy population sample) within the 60-day sample period

We now demonstrate a few real-life scenarios that still achieve this health target and yet satisfy different needs of policy makers and populations. The first scenario we use is a case of ramping vaccination availability, which is a challenge many countries and organizations face today. In this scenario, we choose a constant screening test rate of 6% and highlight all entries in the decision tree that achieves our <5% infection rate target. The representative decision tree for this scenario is shown in Figure7 below where only the combinations of ramping vaccination rate and 6% screening test rate are highlighted from the decision tree of Figure6; all other entries are removed. It is evident from Figure7 that with low vaccination rates (e.g., ≤10%) a higher percentage of the population (≥60%) still needs to continue to wear face masks. As the vaccine availability increases, the face masking mandate can be relaxed gradually until we reach 50% vaccination among the sample population. At this point, wearing face masks is not required at all to achieve that health target. It is also worth noting that with 70% or more vaccination, the screening test rate can drop from 6% to 2% and reduce costs for schools or organizations. This is a good example of how decision makers can utilize the simulation results to achieve their health target.

**Figure7.**
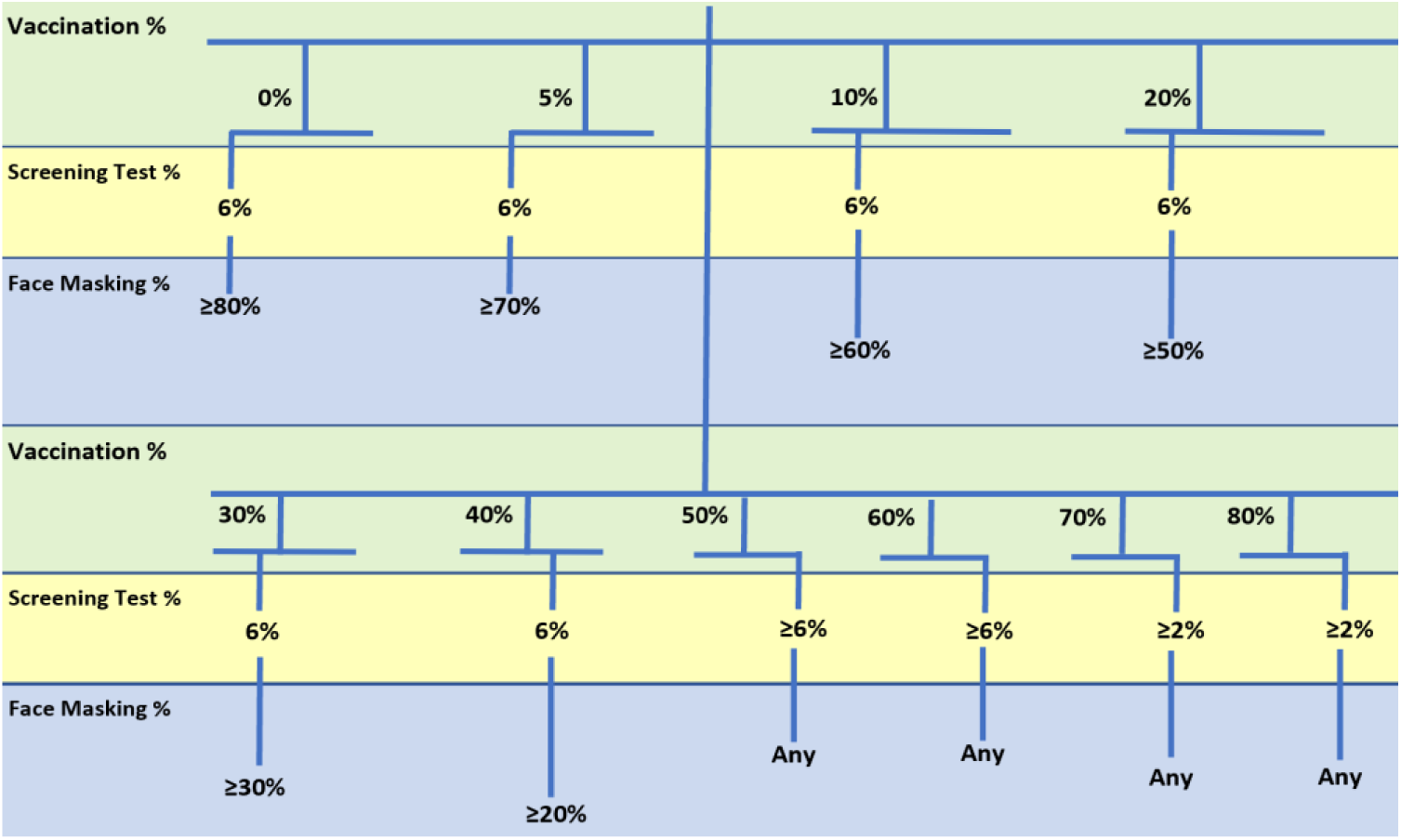
A ramping vaccination rate scenario with an assumed constant screening test rate. All combinations shown achieve the <5% infection rate health target.

To demonstrate another example scenario, we assume policy makers’ goal to eliminate face masking and other social mitigation mandates. The decision tree depicted in Figure8 highlights all combinations that achieve the health goal with no face mask mandate. With lower vaccination rates (≤20%) a high percentage of daily test screening (20%) is needed to achieve the health target goal of <5% infection rate. As the vaccine rate increases, the screening test rate can be reduced gradually until reaching ≥70% vaccination rate, at which screen testing can be as low as 2%. This scenario is applicable to a school population where enforcing face masking is both uncomfortable and unrealistic for young students.

**Figure8.**
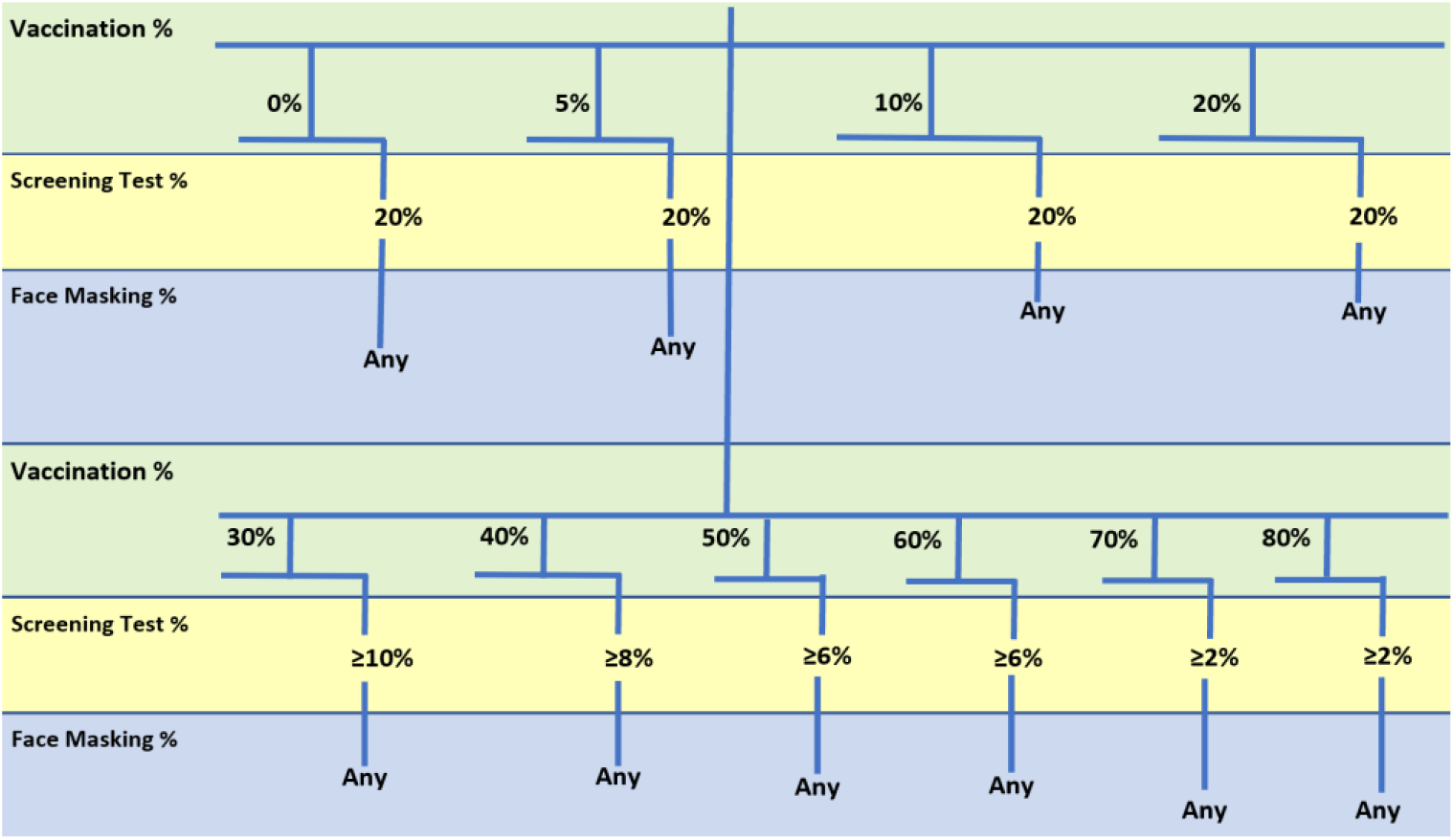
An example scenario of eliminating face masking mandates. All combinations shown achieve the <5% infection rate health target while removing any face masking mandates.

The third scenario we demonstrate is a case where the cost of the daily screening test is a challenge for certain organizations. The cost of daily testing over the course of 60 days or longer adds up quickly as it is a daily cost rather than a one-time cost for each person, like vaccination or face masking. Figure9 demonstrates this cost-sensitive scenario where the minimum screening test percentage is chosen for each corresponding vaccination rate. As shown in the figure, with lower vaccination rates, higher face masking percentages need to be implemented among the population. As the vaccination rate ramps incrementally, the minimum screening test rate decreases while the face masking percentage also decreases. When vaccination rates reach ≥70, the screening test can be completely removed, and its cost can be saved.

**Figure9.**
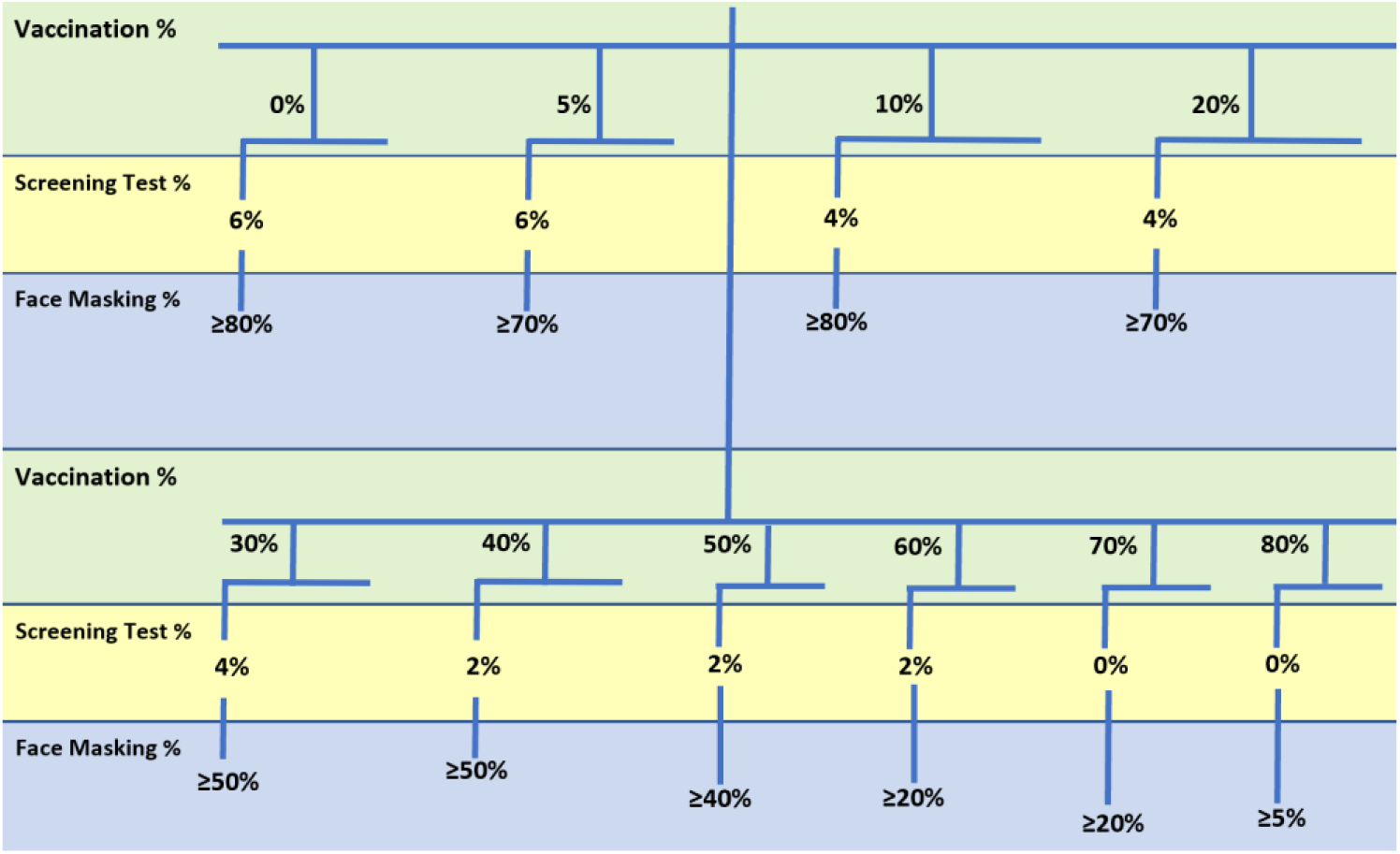
An example scenario of a cost-sensitive decision making where the minimum screening test percentage for each corresponding vaccination rate is chosen to minimize total cost while still achieving the health target. All combinations shown achieve the <5% infection rate health target.

To gain more insights into the simulation results, we analyzed the cases where only two out of the three prevention methods are available. For example, if the vaccine is not yet readily available in certain countries or its cost is prohibitive to certain communities then we need to evaluate how efficient only the screening tests and face masks will be. If the screening test is not applicable to certain organizations, due to incurred cost for example, then it is also important to examine how vaccination and face masking work in tandem to protect the sample population. Finally, face masking may not be a practical option for young students or certain populations who oppose the concept. Table3 below lists the vaccination percentages against the face masking percentages in the case of no screening tests performed (SP=0%.) For completeness purposes, we added a few more simulation combinations when the vaccination rate plus the face masking rate is 100%, e.g., 20% vaccination rate and 80% face masking rate. Since 1% of the population is assumed infected, we used 1% less face masking for these cases (e.g., 79% face masking). The additional simulation combinations are marked in Table3. Figure10a shows a visual representation of this case with a surface plot graph. Figure10b & c demonstrate box plot examples for selected curves on the surface plots where either vaccination is fixed at 30% or face masking is fixed at 30%; the two curves are marked on the surface plot. The box plot shows the median, 25^th^/75^th^ percentiles, the min/max, and the outliers.

**Table3.** Unaffected population % vs. combinations of vaccination % and face masking % with no screening test

| Vaccination % | 0% | 5% | 10% | 20% | 30% | 40% | 50% | 60% | 70% | 80% |
| --- | --- | --- | --- | --- | --- | --- | --- | --- | --- | --- |
| Face Masking % |  |  |  |  |  |  |  |  |  |  |
| 0% | 2 | 5 | 8 | 16 | 25 | 37 | 51 | 67 | 85 | 94 |
| 5% | 2 | 6 | 9 | 17 | 27 | 39 | 52 | 71 | 88 | 95 |
| 10% | 3 | 6 | 11 | 19 | 30 | 43 | 57 | 78 | 91 | 97 |
| 20% | 5 | 8 | 12 | 23 | 36 | 53 | 73 | 90 | 96 | 98* |
| 30% | 6 | 11 | 17 | 29 | 44 | 63 | 83 | 94 | 98* |  |
| 40% | 8 | 14 | 21 | 38 | 54 | 74 | 93 | 96* |  |  |
| 50% | 13 | 19 | 27 | 46 | 67 | 89 | 95* |  |  |  |
| 60% | 18 | 25 | 37 | 70 | 83 | 95* |  |  |  |  |
| 70% | 27 | 36 | 53 | 79 | 91* |  |  |  |  |  |
| 80% | 42 | 54 | 71 | 89* |  |  |  |  |  |  |

\*These data points are additional to the 504 simulation cases. For completeness of the table, we simulated the cases where the face masking percentage is 1% less than the corresponding table value. This was needed to account for the 1% assumed initial infection population. For example, vaccination rate of 20% used 79% face masking rate to add up to 100% with the 1% infection rate.

**Figure10.**
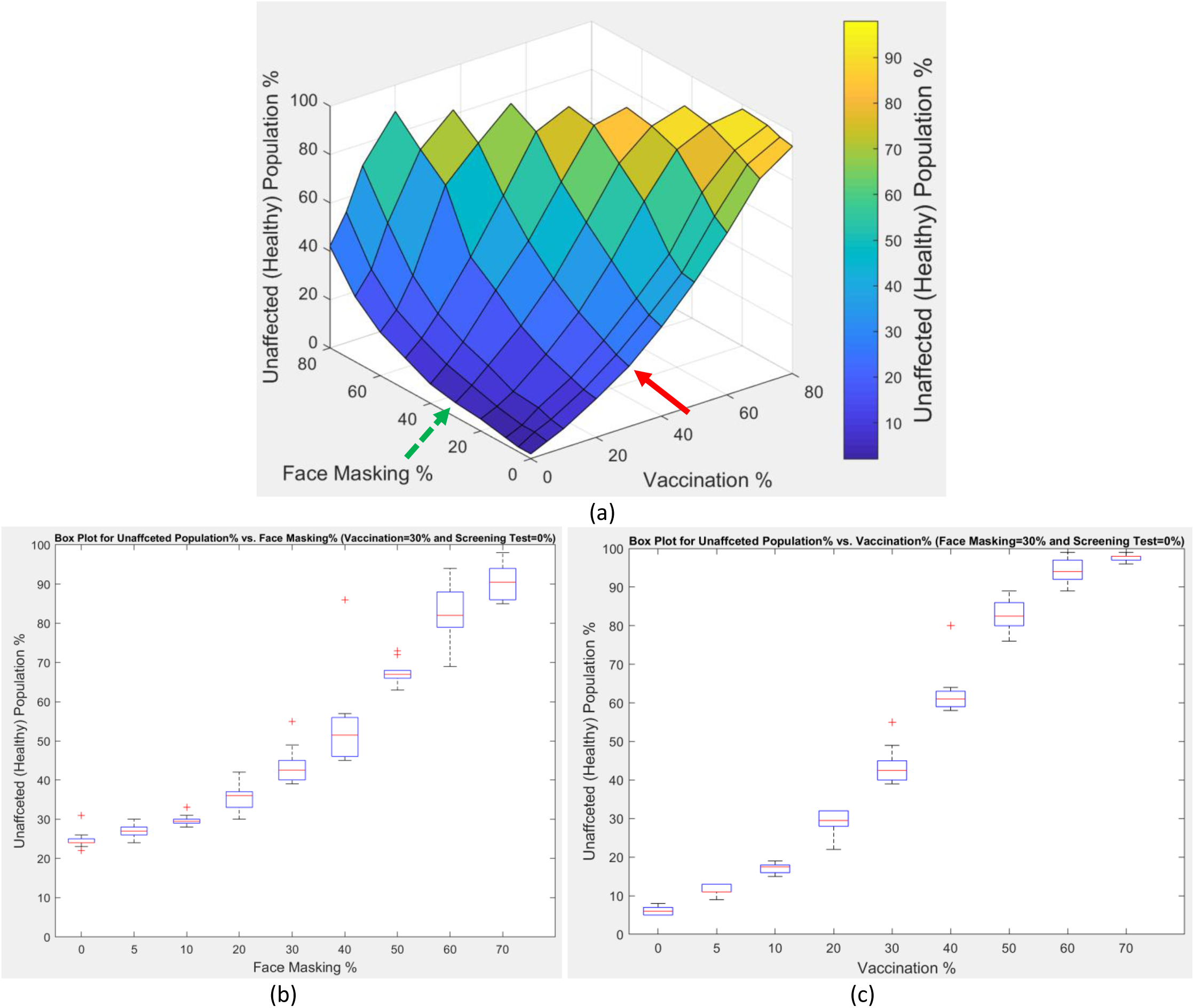
(a) A surface plot for the unaffected (healthy) population % vs. the combinations of the vaccination % and face masking % while there is no screening test applied. (b) A box plot for the unaffected population % vs. face masking % (vaccination=30%, curve marked by the red solid arrow) (c) A box plot for the unaffected population % vs. vaccination % (face masking=30%, curve marked by the green dashed arrow)

Some observations from this data are:

➢ With low vaccination rates, face masking alone will not be sufficient to protect the population and contain the virus infection spread, even with a very high percentage of the sample population abiding by the face masking rules. For example, with vaccination rate of 5%, the protection rate is only 54% with the highest face masking rate of 80% of the sample population.
➢ On the contrary, with low face masking rates, high vaccination rates can help achieve high protection rates. For example, with 5% face masking, the protection rate can reach up to 88% of the population with 70% vaccination, or it can be 95% with 80% vaccination.
➢ By inspecting the surface plot in Figure10a, it is evident that the slope of the healthy population along the vaccination axis is steeper than the slope of the healthy population along the face masking axis, thus indicating that vaccination is a more effective prevention method than face masking in our case.
➢ The box plot in Figure10b demonstrates a case where there is no screening test available while the vaccination rate is still at 30% within the sample population. In this scenario, lower face masking rates result in lower protection. As the face masking rate increases, the protection rate increases slowly, but the protection does not go over 91% even with face masking rate close to 70% in this case.
➢ In contrast, Figure10c box plot graph demonstrates a case where face masking is at 30% and there is no screening test available. In this scenario, the protection rate increases more rapidly while increasing the vaccination rate until it reaches 94% protection with 60% vaccination rate or 98% with 70% vaccination rate, which highlights the importance of vaccination.

In Table4 we list the unaffected population % vs. the combinations of screening test % and face masking % while there is no vaccination applied. Figure11 shows the surface plot for such data, along with two box plots of selected curves on the surface plot to demonstrate the data spread. The following can be inferred from this data:

**Table4.** Unaffected population % vs. combinations of screening test % and face masking % with no vaccination

| Screening Test % | 0% | 2% | 4% | 6% | 8% | 10% | 20% |
| --- | --- | --- | --- | --- | --- | --- | --- |
| Face Masking % |  |  |  |  |  |  |  |
| 0% | 2 | 5 | 17 | 43 | 61 | 87 | 98 |
| 5% | 2 | 7 | 18 | 45 | 72 | 86 | 98 |
| 10% | 3 | 8 | 21 | 47 | 74 | 91 | 97 |
| 20% | 5 | 13 | 41 | 54 | 80 | 94 | 98 |
| 30% | 6 | 18 | 47 | 75 | 94 | 94 | 98 |
| 40% | 8 | 24 | 54 | 78 | 94 | 96 | 98 |
| 50% | 13 | 34 | 68 | 87 | 93 | 97 | 98 |
| 60% | 18 | 44 | 75 | 90 | 94 | 97 | 98 |
| 70% | 27 | 60 | 84 | 94 | 96 | 97 | 99 |
| 80% | 42 | 80 | 91 | 96 | 98 | 98 | 99 |

**Figure11.**
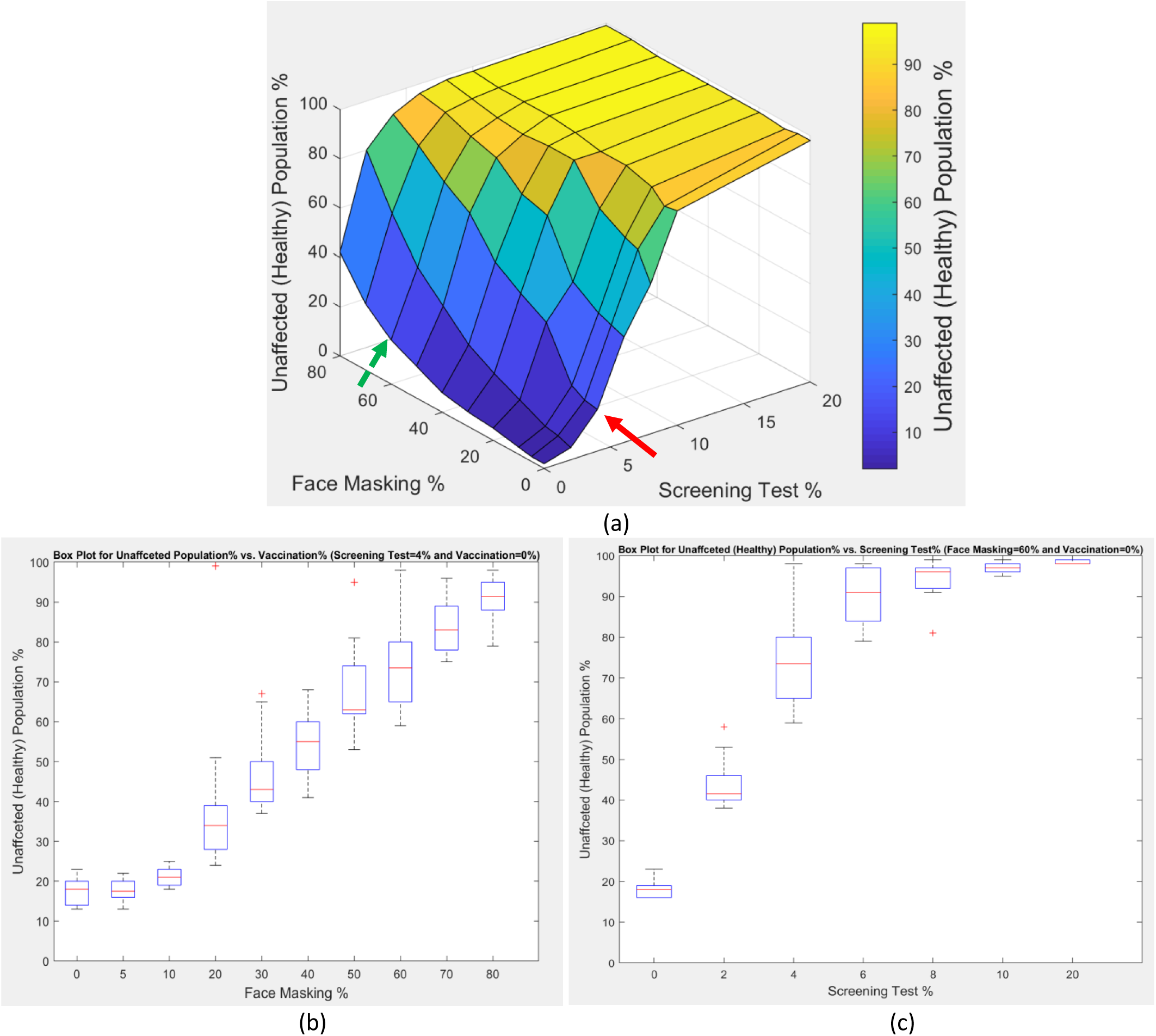
(a) A surface plot for the unaffected (healthy) population % vs. the combinations of the screening test % and face masking % while there is no vaccination applied. (b) A box plot for the unaffected population % vs. face masking % (screening test=4%, curve marked by the red solid arrow) (c) A box plot for the unaffected population % vs. screening test % (face masking=60%, curve marked by the green dashed arrow)

➢ Even with the lack of vaccination in certain countries or communities, we can still achieve high protection rates with many combinations of screening tests and face mask mandates.
➢ Higher screening test rates (≥10%) achieve high protection rates (≥86%) regardless of the face masking rates applied. At a 20% screening test rate, almost all the sample population (≥97%) is protected from the virus spread without any face masking mandates.
➢ Reasonable screening test rates (6% and 8%) can still achieve high protection rates if coupled with higher face masking rates. For example, with 6% screening test and 70% face masking, a 94% protection rate is achieved. An 8% test rate and 30% face masking rate also protects 94% of the sample population.
➢ With most of the sample population (80%) abiding by the face masking rules, high protection rates (91%) can be achieved with a small screening test rate (4%). This use case can demonstrate a low-cost option for protecting the population with a lack of vaccination and a lack of sufficient funds for higher rates of screening tests.
➢ By inspecting the surface plot of Figure11a, we can see how the protection rate rises rapidly with increasing screening test rates, as indicated by the steep slope along the screening test axis. The protection rate almost saturates at screening test rates ≥10%. On the contrary, the protection rate ramps more slowly along the face masking rate axis.
➢ The box plot in Figure11b demonstrates a case where vaccination is not yet available in the community and test screening is fixed at 4%, for cost reasons. In this scenario, ramping face masking rates increases the protection rate gradually until reaching above 90% protection with 80% face masking, which highlights the importance of adopting a mask mandate on a large scale within the community when no vaccination is available.
➢ The box plot in Figure11c demonstrates a case where there is still no vaccination available, and the face masking rate is fixed at 60%. The protection rate rapidly increases with the increasing screening test rate. Protection rates above 90% can be achieved by a 6% test rate. Test rates above 6% will improve protection rates up to 98% at 20% test rate, but this comes at the increased cost of testing a higher percentage of the sample population daily.

The third case of combining two out of the three prevention methods is shown in Table5 and Figure12. In this case we assume no face masking rules are applied and we tabulate and plot the protection rates vs. the combinations of vaccination rates and screening test rates.

**Table5.** Unaffected population % vs. combinations of vaccination % and screening test % with no face masking

| Vaccination % | 0% | 5% | 10% | 20% | 30% | 40% | 50% | 60% | 70% | 80% |
| --- | --- | --- | --- | --- | --- | --- | --- | --- | --- | --- |
| Screening Test % |  |  |  |  |  |  |  |  |  |  |
| 0% | 2 | 5 | 8 | 16 | 25 | 37 | 51 | 67 | 85 | 94 |
| 2% | 5 | 10 | 15 | 23 | 38 | 55 | 67 | 85 | 95 | 97 |
| 4% | 17 | 23 | 34 | 45 | 60 | 80 | 87 | 94 | 95 | 98 |
| 6% | 43 | 44 | 55 | 74 | 85 | 91 | 96 | 98 | 98 | 98 |
| 8% | 61 | 74 | 73 | 91 | 92 | 97 | 97 | 98 | 98 | 99 |
| 10% | 87 | 88 | 94 | 91 | 95 | 97 | 97 | 98 | 98 | 99 |
| 20% | 98 | 97 | 98 | 98 | 98 | 98 | 98 | 99 | 99 | 99 |

**Figure12.**
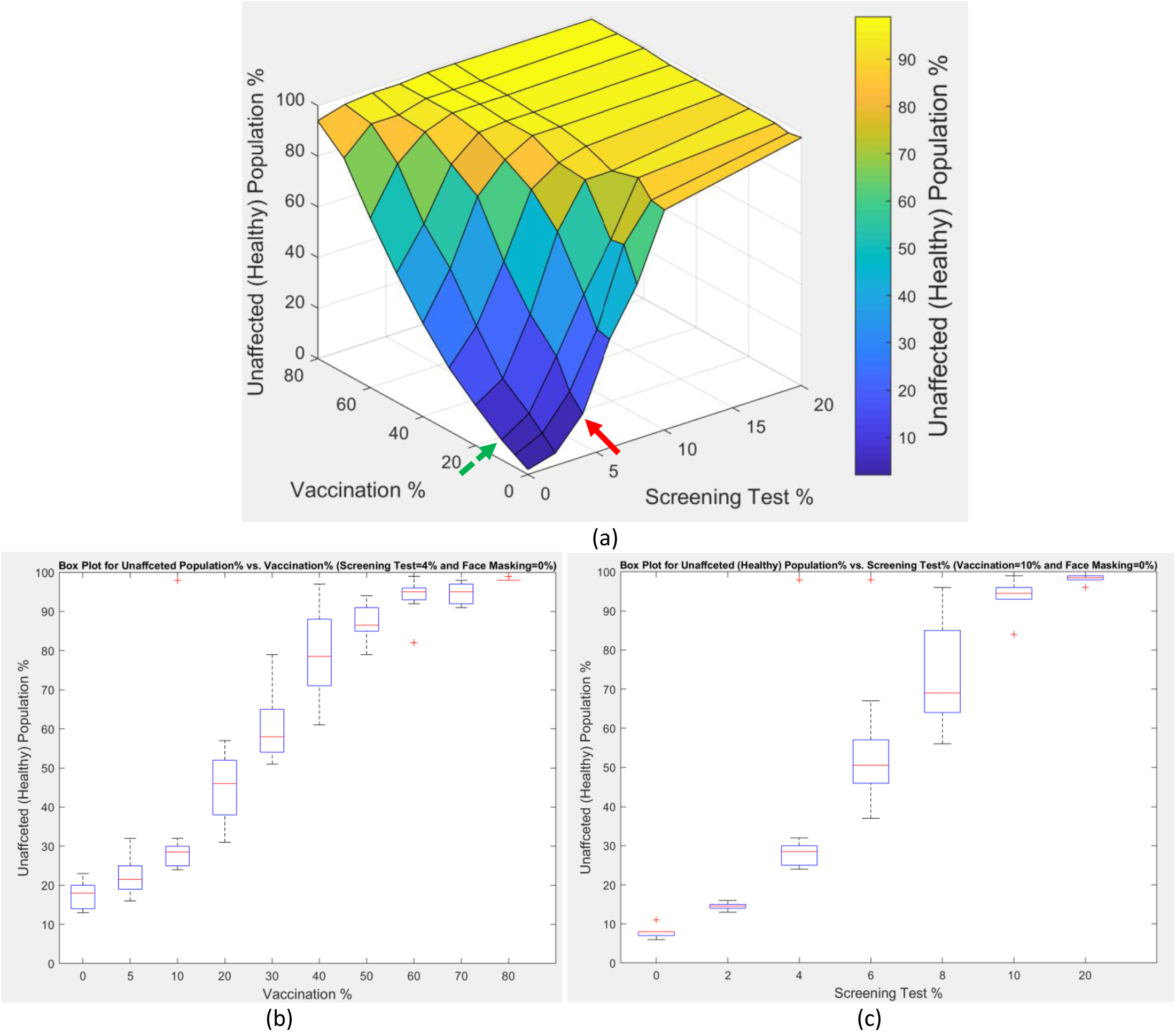
(a) A surface plot for the unaffected (healthy) population % vs. the combinations of the vaccination % and screening test % while face masking is 0% (b) A box plot for the unaffected population % vs. vaccination % (screening test=4%, curve marked by the red solid arrow) (c) A box plot for the unaffected population % vs. screening test % (vaccination=10%, curve marked by the green dashed arrow)

We can infer the following from this data:

➢ These two prevention methods are both highly effective in achieving high protection rates for the sample population under study. This is evident from the many table entries that exceed 90% protection rate for example. Each method alone can achieve the highest protection rates (e.g., 80% vaccination rate alone or 20% screening test rate alone can achieve ≥94% protection) but will achieve it much faster if combined with the other method (e.g., 50% vaccination and 6% screening achieve 96% protection, or 70% vaccination and 2% screening achieve 95% protection). This observation is also visible in the surface plot in Figure12a where the protection rate ramps rapidly along the two axes and very high rates are achieved with reasonable combinations.
➢ This result is crucial to help policy and decision makers to alleviate the face masking mandate if enough population get vaccinated and if a small percentage of the population who can’t or won’t get vaccinated abide by recurring screening test policy.
➢ The box plot in Figure12b demonstrates a case where face masking mandate is completely removed or avoided, and screening test rate is fixed at 4% to reduce cost. In this scenario, as the vaccination rate ramps up, the protection rate increases rapidly until it reaches 94% or above with just 60% or more vaccination. This scenario highlights the importance of vaccination in increasing the protection for the entire population.
➢ The box plot in Figure12c demonstrates a case with no face masking mandate again and a fixed small vaccination percentage of 10%. The protection rate increases rapidly with the increasing screening test rate until it reaches 94% or above protection with 10% or greater test rate. This example highlights the importance of screening tests when the vaccination is not readily available yet and face masking mandate is challenging to apply in the community.

## Conclusions

In this simulation study, we investigated the interplay of the three available SARS-CoV-2 virus spread prevention methods (screening test, vaccination, and face masking) on a small sample population in a confined space, based on some reasonable assumptions for virus transmissibility rates. We demonstrated the efficacy of each method alone and the efficacy of combining two or three methods together. The simulation results should not be taken in an absolute sense as many of the underlying assumptions can be changed, such as the population sample size or the different virus transmissibility rates. However, the observed patterns should still be valid to a greater extent.

Using the decision tree methodology to interpret the results should be a powerful and flexible tool to allow decision makers all over the world to define many scenarios that can fit their environment, timeline, and resources. The three scenarios we demonstrated (ramping vaccination scenario, alleviation of all mask mandates scenario, and cost-sensitive scenario) are all reasonable scenarios that can apply to many places in the world. Many more scenarios can be derived from the simulation data and the decision tree built.

We aim the results of this study to guide policy makers and business leaders on how to combat the virus infection spread. The results should inform decision makers to help open our schools and businesses safely if some of the mitigation scenarios here are implemented. The different combinations that achieve the same health target also provide a lot of flexibility in protecting different populations depending on the demographic, financial, and political conditions for each organization or country. For example, certain developing countries may lack access to vaccination or cannot afford to offer it to the whole population, so relying on combining a screening test strategy with imposing some social mitigation rules like face masking can achieve the target health goal. As vaccination becomes increasingly available, some of the mitigation rules and/or screening tests can be relaxed.

## Data Availability

All data produced in the present study are available upon reasonable request to the authors

## Notes

### Competing Interest Statement

The authors have declared no competing interest.

### Funding Statement

This study did not receive any funding

